# Program Evaluation of the WASHmobile PICHA7 mHealth and Chlorine E-Voucher Program in the Democratic Republic of the Congo

**DOI:** 10.1101/2025.08.31.25334790

**Authors:** Christine Marie George, Jean-Claude Bisimwa, Kelly Endres, Justin Bengehya, Jean-Claude Kulondwa, Raissa Boroto, Ghislain Maheshe, Cirhuza Cikomola, Presence Sanvura

## Abstract

**Background:** Targeted water treatment and hygiene (WASH) programs for those residing near to diarrhea patients can serve as a cost-effective approach during outbreaks to reduce the spread of diarrhea by targeting those at highest risk. Our research team designed the WASHmobile mobile health (mHealth) program for those at high risk of diarrhea. In our randomized controlled trials in the Democratic Republic of the Congo (DRC) (PICHA7) and Bangladesh (CHoBI7), delivery of WASHmobile to diarrhea patient households through a healthcare facility visit and voice and SMS messages from a doctor significantly reduced diarrhea and improved child growth.

**Methods:** Building on this work, we adapted WASHmobile to serve millions more beneficiaries through a mHealth and e-voucher program delivered in diarrhea outbreak areas. A program evaluation of this approach was conducted in health areas (health administrative unit) with ongoing diarrhea outbreaks in the DRC among 2022 participants. Voice and SMS messages were sent to those within 500 meters of diarrhea patients from a doctor stating that there was a diarrhea outbreak in their health area and emphasized the importance of treating and safely storing household drinking water and washing handwashing with soap for the next 7-day high-risk period. The SMS messages contained an e-voucher to redeem for free chlorine tablets at a pharmacy. Unannounced spot checks assessed WASH behaviors a week after program initiation.

**Results:** Fifty-seven percent of WASHmobile households redeemed e-vouchers for chlorine tablets at pharmacies. Compared to control households, WASHmobile households that redeemed e-vouchers had higher stored drinking water with free chlorine concentrations >0.2 mg/L (Odds Ratio [OR]: 6.93, [95% Confidence Interval [CI]: 1.76, 27.24]) (64% [WASHmobile] vs. 20% [control]) and stored drinking water completely covered (OR: 4.55, 95% CI: 2.68, 7.70) (73% vs. 38%). Presence of a cleansing agent within 10 steps of latrine and cooking areas was also significantly higher in WASHmobile households (latrine: OR: 3.64, 95% CI: 1.47, 9.02 [70% vs. 39%] and cooking: OR: 2.50, 95% CI: 1.31, 4.77 [70% vs. 49%]).

**Conclusions:** The WASHmobile PICHA7 mHealth and e-voucher program significantly increased water treatment, safe water storage, and hygiene behaviors in diarrhea outbreak areas in the DRC.

## Introduction

Diarrhea from inadequate water, sanitation, and hygiene (WASH) is estimated to contribute to 54 million disability-adjusted life-years (DALYs) and 1 million deaths globally each year.^1,2^ In the Democratic Republic of the Congo (DRC), diarrhea accounts for 2.3 million DALYs and 2 million inpatient diarrhea cases annually.^1,3^ Chronic diarrhea episodes in children <5 years contribute to undernutrition, increasing the risk of stunting, which is associated with mortality and impaired cognitive development.^4-7^ Furthermore, climate change has increased droughts and floods in Africa, driving diarrhea outbreaks to historic highs in the region.^8,9^ These outbreaks burden healthcare systems in low- and middle-income countries worldwide.^1^

Early alert and response systems for diarrhea outbreaks that target populations at highest diarrhea risk promote climate resilience and are a critical cost-saving measure to reduce diarrhea morbidity and mortality compared to a blanket approach. Our research in the DRC has shown that when a diarrhea patient is admitted to a healthcare facility, individuals living within 500 meters are at >12 times higher risk of hospitalized diarrhea than the general population for the following 7 days (George et al. submitted). Targeted WASH programs for those residing near diarrhea patients therefore represent a cost-effective approach during outbreaks to reduce the spread of diarrhea by targeting those at highest risk. This type of targeted strategy builds on the status quo of waiting until diarrhea outbreaks become large before responding with a resource-intensive “blanket approach,” where WASH interventions are delivered broadly in-person to large population which is costly and time-consuming.^10^

In partnership with the Ministries of Health in the DRC and Bangladesh, we developed a WASH mHealth messaging system for diarrhea patient households as part of the WASHmobile program.^11,12^ This approach was evaluated among diarrhea patient households in two recent randomized controlled trials (RCTs) of WASHmobile: in the DRC (PICHA7 site, N=2,334 participants) (current site)^13^ and in Bangladesh (CHoBI7 site, N=2,626 participants).^14^ In these RCTs, we found that sending diarrhea patient households weekly automated WASH-related voice and SMS mHealth messages from a doctor, along with provision of chlorine tablets and soapy water (water and detergent powder) resulted in significantly higher sustained water treatment and handwashing with soap, as well as significant reductions in healthcare visits for diarrhea, diarrhea prevalence, and stunting over a 12-month period.^13,14^

Building on this work, in partnership with the DRC Ministry of Health, we have now adapted the WASHmobile PICHA7 program to serve millions more beneficiaries through a location-based push notification system. The system delivers WASH mHealth messages and e-vouchers for chlorine tablets to redeem at local pharmacies to those in high-risk areas for diarrhea outbreaks. This program is delivered in health areas (a DRC government health administrative unit of ∼10,000 individuals) at the beginning stages of diarrhea outbreaks. Voice and SMS messages are sent to those within 500 meters of diarrhea patients from a doctor stating that there is a diarrhea outbreak in their health area, emphasizing the importance of treating and safely storing household drinking water and washing hands with soap for the next 7-day high-risk period. The SMS messages also contain an e-voucher to redeem for free chlorine tablets at a local pharmacy.

In this study, we investigated whether delivery of the WASHmobile PICHA7 mHealth and chlorine E-Voucher program can increase water treatment, safe water storage, and hygiene behaviors among those living within 500 meters of diarrhea patients during diarrhea outbreaks.

## Methods

This study conducted a program evaluation of the WASHmobile PICHA7 mHealth and chlorine E-Voucher program implemented between December 2024 and July 2025 during ongoing diarrhea outbreaks in DRC. Evaluation activities were conducted in Bukavu, a city with a population of >1 million in eastern DRC. Evaluation activities were unannounced spot checks of water treatment, safe water storage, and hygiene indicators. A “diarrhea outbreak health area” is defined at our study site as a health area where there are ≥5 inpatient diarrhea cases residing in the same health area admitted to a healthcare facility for treatment within 24 hours. This alert of a diarrhea outbreak health area is based on the location where patients report residing (not the location where patients sought care). Inpatient diarrhea cases are captured through daily surveillance at 115 public and private healthcare facilities in Bukavu conducted in partnership with the DRC Ministry of Health.^15^

### WASHmobile PICHA7 mHealth and Chlorine E-Voucher program

The WASHmobile PICHA7 mHealth and chlorine e-voucher program builds on our previous program through: (1) broadening the scope of the program from focusing on diarrhea patient households to including those residing in diarrhea outbreak health areas, allowing the program to serve millions more beneficiaries; and (2) leveraging new technologies: (i) location-based push notifications; and (ii) chlorine e-vouchers to be redeemed at local pharmacies. This program is delivered in health areas at the beginning stages of diarrhea outbreaks with a diarrhea outbreak health area alert.

The WASHmobile PICHA7 mHealth and chlorine e-voucher program includes the delivery of three automated voice calls and three SMS messages from a doctor at the provincial hospital during the 7-day high-risk period after the diarrhea outbreak alert in the health area. All mHealth messages were reviewed and recorded by author RB, a physician in the cholera treatment ward at the Provincial Hospital who sent messages under the name Dr. Picha to align with the name of the intervention program. Mobile messages include interactive voice response (IVR), voice, and SMS messages. For IVR messages, participants are asked to respond 1 or 2 in response to a question, after which they receive a response with the correct answer. There is no charge to the phone subscriber receiving IVR or voice calls or SMS messages. The web-based engageSPARK platform is used to send program messages.^16^ When a diarrhea outbreak health area is identified, a health worker collects the phone number of the inpatient diarrhea case at the healthcare facilities where patients are identified and contacts the community health worker in their health area to collect the GPS location of the patient household. In the DRC, public community health workers are embedded in the health system at the health area level making this approach feasible to deliver at scale. Within 48 hours of a diarrhea outbreak alert in the health area, a voice and SMS message from the WASHmobile PICHA7 mHealth and e-voucher program are sent to all phone subscribers within an approximately 500-meter radius of the diarrhea patient. This can be done either through location-based push notification sent by mobile network operators or by the community health worker collecting the numbers of individuals residing within 500 meters of the diarrhea patient household. Messages are sent in the evening at 8 PM when most individuals are home.

The SMS message contains a unique chlorine e-voucher ID (e.g., 6DE78-86518) and the location of a designated pharmacy in the phone subscriber’s health area where the e-voucher can be redeemed for chlorine tablets. Commonly used pharmacies in health areas that supplied chlorine tablets were selected. Phone subscribers have 7 days from the time the SMS message is sent to bring this unique e-voucher number (either on their phone or written down) to their designated pharmacies to redeem for 8 chlorine tablets (160 liters of treated water). When phone subscribers bring the e-voucher number to the pharmacy, the pharmacy employee enters a short code into their phone with the unique e-voucher ID and they receive a reply back on whether the e-voucher is valid (e.g., unexpired and unused). No smart phone capability is needed for e-voucher validation, a feature phone can be used. If the e-voucher ID is valid, phone subscribers are provided the chlorine tablets. The date and time the e-voucher was redeemed is recorded through the database designed on Kobo Toolbox. After the 7-day high risk period in the diarrhea outbreak health area is complete, pharmacy staff receive a stipend for participation in the program and reimbursement for all e-vouchers redeemed. An example voice, IVR, and SMS message from the WASHmobile PICHA7 program are shown here.

***WASHmobile Voice Call***

*Hello. I am Dr. Picha calling from Provincial Hospital of Bukavu. There is a diarrhea outbreak in the Brasserie health area. Many people are coming to our hospital for diarrhea treatment. Protect yourself and your family from diarrhea by washing your hands with soap before eating and after toileting over the next 7 days. In addition, you should treat your water with chlorine tablets and always completely cover your stored water. We are sending you a SMS with a voucher for free chlorine tablets to redeem at Biosadec Pharmacy located in Brasserie, opposite the market where the wooden planks are sold. Bring this SMS to Biosadec Pharmacy in the next 7 days to receive free chlorine tablets. We will send you messages over the next week with updates*.

***WASHmobile SMS***

*This is Dr. Picha from Provincial Hospital of Bukavu. There is a diarrhea outbreak in Brasserie health area. Many people are coming to our hospital for diarrhea treatment. Protect yourself and your family from diarrhea by using chlorine tablets for your drinking water, completely covering stored water, and washing your hands with soap over the next 7 days. Bring your phone with this SMS e-voucher number 6DE78-86518 to Biosadec Pharmacy, located in Brasserie, opposite the market which sells wooden planks to redeem your free chlorine tablets. You have the next 7 days to redeem these chlorine tablets. -Dr. Picha*

***WASHmobile IVR***

*Hello! This is Dr. Picha calling from the Provincial Hospital of Bukavu to remind you that there is a diarrhea outbreak in your health area. Many people are coming to our hospital from your health area for diarrhea treatment. Please listen to my important information. You should have received an e-voucher by SMS for free chlorine tablets to redeem at our local pharmacy. You have 7 days to bring this e-voucher to Biosadec Pharmacy to receive these free chlorine tablets. This pharmacy is located in Brasserie, opposite the market where the wooden planks are sold. These chlorine tablets are important to protect you and your family from severe diarrhea. Now I want to ask you a question. Please answer by pressing 1 or 2 on your phone. There is no cost for responding*.

*How should you treat your drinking water using chlorine tablets?*

*If you think you should add one chlorine tablet to a 20-liter jerry can and wait 30 minutes before drinking press 1*.

*If you think you should add one chlorine tablet to a 20-liter jerry can and wait 60 minutes before drinking press 2*.

### Study Design for Program Evaluation

The WASHmobile PICHA7 mHealth and chlorine e-voucher program was delivered in two Bukavu health areas (Irambo and Burhiba) to those phone subscribers within 500 meters of diarrhea patients. Diarrhea patients were screened at 115 public and private healthcare facilities surveillance healthcare facilities. In control areas, we enrolled 10 diarrhea patients in non-diarrhea outbreak health areas and 10-12 households within 500 meters of each of these patients. Households were enrolled and spot checks were conducted within 7 days of the alert of a diarrhea outbreak in the health area. Unannounced spot checks were conducted in WASHmobile PICHA7 program and control households. Spot checks were unannounced to prevent households preparing for our arrival. After enrollment, a household stored drinking water sample was collected to test for free chlorine. Chlorine was measured using a digital colorimeter (Hach, Loveland, CO, USA). The World Health Organization (WHO) guideline for free chlorine in household stored drinking water of >0.2 mg/L was used as the cutoff for chlorination.^17^ During unannounced spot checks, the covering status of household stored drinking water (proxy indicator of safe water storage behaviors) and the presence of a cleansing agent (bar soap, liquid soap, soapy water, or ash in a container) and water within 10 steps of the latrine and cooking area (proxy indicator of handwashing with soap behavior) were observed.^18^ Information was also collected on the type of drinking water storage container and on household water source type using the categories defined by the WHO/ UNICEF Joint Monitoring Program.^19,20^ Process evaluation indicators of mHealth message delivery were collected using the engageSPARK platform on the percentage of unique voice, IVR, and SMS messages received by program households.

### Statistical Analysis

The sample size was based on the number of program and control areas that could be enrolled given security considerations due to ongoing armed conflict in the area. To evaluate the effectiveness of the WASHmobile PICHA7 program compared to control areas, logistic regression models were performed to compare WASH outcomes by area type (WASHmobile or control) using general estimating equations (GEE) to account for clustering in the health area and to approximate 95% confidence intervals. To compare water source type, drinking water container type, and the presence of oral rehydration solution in the home we performed a fisher exact test due to sample size constraints. Statistical analyses were conducted using SAS 9.4 software (Cary, NC, USA).

### Ethical approval

Ethical approval was obtained from the institutional review boards of the Johns Hopkins School of Public Health and Catholic University of Bukavu. Written informed consent was obtained from all individuals or their guardians participating in data collection activities.

## Results

Program evaluation activities assessed WASH and mHealth outcomes among 2022 participants, 872 participants in diarrhea outbreak health areas receiving the WASHmobile PICHA7 mHealth and chlorine e-voucher program (132 households) and 1150 participants (157 households) in control areas (Table 1). Enrolled participants resided in a total of nine health areas. Demographic characteristics are reported in Supplementary Table 1.

**Table 1.** Study population and PICHA7 mHealth intervention received

|  | PICHA7 E-Voucher Households |  |  | Control Households |  |  |
| --- | --- | --- | --- | --- | --- | --- |
|  | % | n | N | % | n | N |
| Total Participants |  |  | 872 |  |  | 1150 |
| Participants per household (median ± sd (range)) | | 6 ± 2.53 (1-15) | | | 7 ± 2.89 (2-16) | |
| Households |  |  | 132 |  |  | 157 |
| PICHA7 mHealth messages received |  |  |  | - | - | - |
| Voice messages | 99% | 271 | 274 | - | - | - |
| Per household (median ± sd (range)) | | 2 ± 0.15 (1-2) | | - | - | - |
| IVR messages | 97% | 411 | 422 | - | - | - |
| Per household (median ± sd (range)) | | 3 ± 0.58 (1-4) | | - | - | - |
| Text messages | 89% | 608 | 686 | - | - | - |
| Per household (median ± sd (range)) | | 5 ± 0.09 (5-6) | | - | - | - |
| Chlorine E-voucher received | 100% | 132 | 132 | - | - | - |
| Chlorine E-voucher redeemed | 57% | 75 | 132 | - | - | - |

SD: standard deviation; IVR: interactive voice response; Received: unique messages received (answered for voice and ivr)

Ninety-nine percent of unique voice messages (271/274) and 97% (411/422) of IVR messages sent to WASHmobile households were answered by households (messages received: voice calls 2 [range: 1-2] and IVR calls: 3]range:1-4]). Eighty-nine percent (608/686) of unique SMS messages were received (mean number of SMS received: 6 [range: 5-6]). All WASHmobile households received the SMS chlorine e-voucher at least once (N=132). Fifty-seven percent of households (75/132) that received chlorine e-vouchers redeemed them for chlorine tablets at their designated pharmacies.

Compared to control households, stored drinking water free chlorine >0.2 mg was significantly higher in WASHmobile households during unannounced spot checks (Odds Ratio [OR]: 6.93, 95% Confidence Interval [CI]: 1.76, 25.24) (64% [WASHmobile] vs. 20% [control]) (Table 2). Household stored drinking water being completely covered was also significantly higher in WASHmobile households compared to control areas (OR: 4.55, 95% CI: 2.68, 7.70) (73% vs. 38%).

**Table 2.**
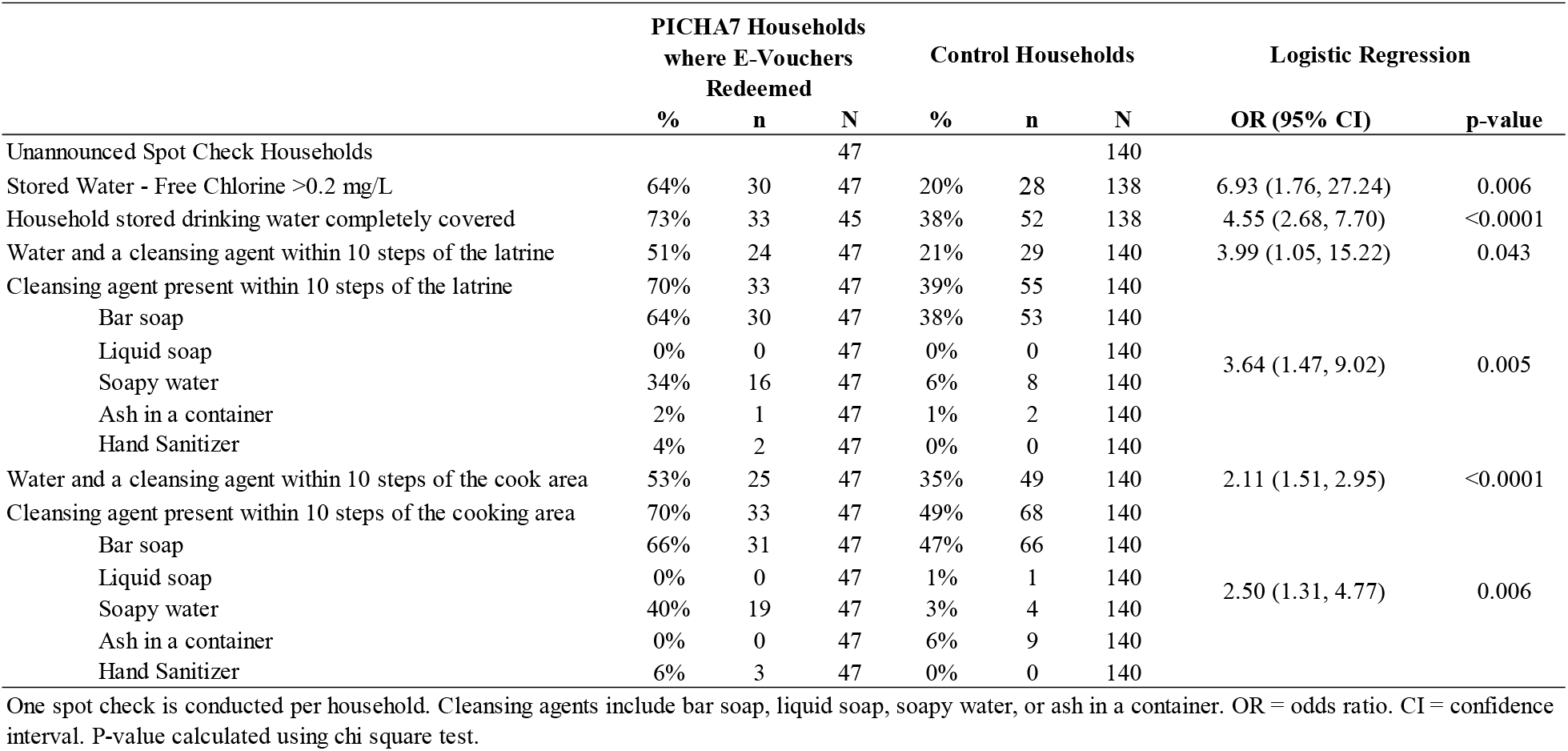
Unannounced spot checks of WASH indicators comparing PICHA7 Households where E-Vouchers were redeemed to control households

For hygiene indicators, presence of a cleansing agent within 10 steps of latrine and cooking areas was significantly higher in WASHmobile households compared to control households (latrine areas: OR: 3.64, 95% CI: 1.47, 9.02 [70% vs. 39%] and cooking areas: OR: 2.50, 95% CI: 1.31, 4.77 [70% vs. 49%]). Consistent with this, presence of water *and* a cleansing agent was also significantly higher within 10 steps of latrine and cooking areas in WASHmobile households compared to control households (latrine areas: OR: 3.99, 95% CI: 1.05, 15.22 [51% vs. 21%] and cooking areas: OR: 2.11, 95% CI: 1.51, 2.95 [53% vs. 35%]). Bar soap was the most common cleansing agent within 10 steps of latrine (64% [WASHmobile] vs. 38% [control] and cooking areas (66% vs. 47%).

## Discussion

Delivery of the WASHmobile PICHA7 mHealth and chlorine e-voucher program in diarrhea outbreak areas resulted in the majority of households redeeming e-vouchers for chlorine tablets at local pharmacies. Furthermore, most of the households redeeming e-vouchers had chlorine in stored drinking water >0.2 mg/L, stored drinking water completely covered, and a cleansing agent in cooking and latrine areas; all of which were significantly higher than control households. This is the first study we are aware that has used e-vouchers for chlorine tablet distribution. Through this study, we were able to demonstrate the technological feasibility and acceptability of e-vouchers redemption for WASH products in the DRC. These findings suggest that the WASHmobile PICHA7 program, which combines mHealth messaging with e-vouchers for chlorine tablets in diarrhea outbreak areas, presents a promising approach to increase water treatment, safe water storage, and proper hygiene practices during diarrhea outbreaks in the DRC.

The high voucher redemption rate and water treatment adherence observed with our WASHmobile PICHA7 program is consistent with two previous RCTs conducted in Malawi and Kenya which used paper vouchers.^21,22^ An RCT in Malawi found that a higher proportion of households that received monthly paper vouchers for free chlorine had chlorine present in stored drinking water compared to control households receiving free chlorine distribution directly through community health workers home visits.^22^ Additionally, paper vouchers for chlorine significantly reduced diarrhea among children compared to control households, with no observed impact for households receiving chlorine directly through community health workers home visits. In Kenya, an earlier RCT found that monthly paper vouchers for free chlorine that could be redeemed at a shop or clinic led to similar rates of chlorine presence in stored household drinking water to chlorine distribution through home or clinic visits (35% vs. 34%).^21^ These promising results demonstrate the positive impact of chlorine vouchers on water treatment behaviors. Future studies are needed that test our WASHmobile mHealth and chlorine e-voucher service delivery model in different contexts globally and compare this to the status quo of blanket door-to-door chlorine product distribution programs.

Targeted delivery of WASH mHealth messages and chlorine e-vouchers to high-risk populations for diarrhea outbreaks has several advantages over the status quo approach of blanket door-to-door community-based WASH programs. First, this approach reduces wastage by targeting households that will likely benefit the most from the use of chlorine products. Previous studies have shown that chlorination rates are similar when chlorine is provided through vouchers compared to free distribution.^21,22^ However, free door to door distribution leads to a much larger number of chlorine products being handed out that may ultimately go unused. Redeeming vouchers comes with a “hassle cost,” since households must take the time to travel to pharmacies to exchange them for chlorine tablets.^21,22^ As a result, households that are not truly interested in using chlorine products are less likely to redeem vouchers, even though they might have accepted the products if they were delivered directly to their homes. Second, delivering mobile messages with e-vouchers through automated, location-based push notifications from mobile network operators to individuals within 500 meters of patients reduces the burden for already overextended community health workers, who would overwise need to make door-to-door visits, and reduces program costs. The bulk cost of mHealth message delivery for the WASHmobile mHealth and chlorine e-voucher program at our site in the DRC is 0.32 USD for 3 voice calls and 3 SMS (no charge to mobile user), and 0.40 USD for an SMS e-voucher for 8 free chlorine tablets (160 L treated water) to redeem at a pharmacy or shop (including a stipend for the pharmacist). Third, the e-voucher system requires households to come to pharmacies or shops, which has the potential to increase market demand for chlorine products and strengthen chlorine product supply chains, reducing stockouts.

Beyond its impact on chlorination of stored household drinking water, the WASHmobile PICHA7 mHealth and chlorine e-voucher program also significantly increased covering of stored drinking water and the presence of a cleansing agent at household latrine and cooking areas. The WASHmobile program included IVR, voice, and SMS messages on safe water storage and handwashing with a cleansing agent at food and stool related events. These promising findings demonstrate the effectiveness of this program on safe water storage and hygiene behaviors in the household even without the provision of bar soap and a water storage vessel. Future studies should assess the impact of this intervention approach on handwashing with soap behavior assessed through structured observation among both program and control households.

Future studies should explore the reasons for households not redeeming chlorine e-vouchers and for the lack of chlorine use among some of the households that have redeemed e-vouchers. Based on our previous formative research and the observations of the study team, potential reasons households did not redeem e-vouchers include distance to the pharmacy, lack of time to pick up chlorine at pharmacies, dislike of the taste of chlorine in drinking water, and low perceived susceptibility to diarrhea (e.g., because no one was ill with diarrhea in their household and/or the belief that chlorine tablets were a treatment for diarrhea).^11,23^ Potential reasons participants redeemed e-vouchers but did not use them include insufficient quantity of water for treatment (need at least 20 liters of water needed) or saving tablets for use when someone in their household had diarrhea.^21,22^ In 2016, Dupas et al. found a negative association between household wealth and redemption of chlorine vouchers, concluding that this relationship was likely because richer household have a higher value of time.^21^ This association should be explored for redemption of e-vouchers in a future study.

The service delivery model for WASHmobile must be tailored to the context for which the program will be implemented. For example, rural and internally displaced people (IDP) camps may vary from urban contexts on factors such as market availability of chlorine products and cellular network connectivity. A market analysis of supply chains and suppliers of chlorine products should be performed prior to implementing this program to ensure chlorine product availability and to prevent stockouts. In remote rural areas and some IDP contexts with low cellular network coverage and mobile phone ownership, paper vouchers will likely be needed to complement mobile messages and e-vouchers for those without cellular access. Community health workers will likely also need to be engaged in WASH promotion and providing paper vouchers for these households. Furthermore, the type of healthcare facilities where diarrhea patients seek treatment will vary between urban, rural, peri-urban, and IDP contexts. Therefore, it will be important that diarrhea patient surveillance includes the different types of healthcare facilities where patients seek treatment to ensure diarrhea outbreaks can be detected early and controlled.

This study has a few limitations. First, we did not assess health outcomes such as reported diarrhea or assess enteric pathogens such as cholera. Health outcomes should be included in future studies. Second, we did not follow households longitudinally to assess whether the intervention resulted in sustained water treatment practices over time. This should additionally be investigated in future studies. Finally, our study only focused on an urban context. Future studies should evaluate WASHmobile delivery in rural, IDP camp, and peri-urban contexts.

This study has several strengths. First, the inclusion of a control area where diarrhea patients resided but did not receive the intervention, building on traditional before and after comparison for program evaluations without a control area. Second, the study included process evaluation data on the number of mobile messages received in addition to chlorine e-vouchers redeemed. This allowed us to determine whether households were answering program voice and IVR calls and receiving SMS messages. Third, unannounced spot checks were conducted to assess the covering status of household stored drinking water and the presence of a cleansing agent and water in household cooking and latrine areas. This builds on previous studies that typically only include chlorine presence in stored drinking water.

The WASHmobile PICHA7 mHealth and e-voucher program significantly increased water treatment, safe water storage, and hygiene behaviors in diarrhea outbreak areas in the DRC. Furthermore, our findings demonstrate the technological feasibility and acceptability of delivery of WASH mHealth messages and chlorine e-vouchers to those in diarrhea outbreak areas in an urban context in the DRC. These findings suggest that the WASHmobile PICHA7 mHealth and e-voucher program presents a promising, low-cost service delivery model to increase WASH behaviors during diarrhea outbreaks. We are currently partnering with the DRC Ministry of Health to develop strategies to scale this program in the DRC through the National Cholera Control Plan.

## Data Availability

All data produced in the present study are available upon reasonable request to the authors.

---

**Supplementary Table 1.** Household Water Source Type and Quality

|  | PICHA7 Households<br>where E-Vouchers<br>Redeemed |  |  | Control Households |  |  | p-value* |
| --- | --- | --- | --- | --- | --- | --- | --- |
|  | % | n | N | % | n | N |  |
| Unannounced Spot Check Households |  |  | 47 |  |  | 140 |  |
| Water Source Type |  |  | 47 |  |  | 136 |  |
| Basic Water Source | 79% | 37 | 47 | 74% | 100 | 136 | 0.74 |
| Limited Water Source | 19% | 9 | 47 | 14% | 19 | 136 |  |
| Unimproved Water Source | 0% | 0 | 47 | 1% | 1 | 136 |  |
| Other | 11% | 1 | 47 | 12% | 16 | 136 |  |
| Drinking Water Container Type |  |  | 45 |  |  | 136 |  |
| Plastic Jerry Can | 84% | 38 | 45 | 97% | 132 | 138 | 0.002 |
| Bucket | 13% | 6 | 45 | 1% | 2 | 138 |  |
| Other | 2% | 1 | 45 | 3% | 4 | 138 |  |
| Days since intervention delivery for spot checks (median) | 4 ± 1.73 (2-10) |  |  | - | - | - |  |

\* p-value calculated with Fisher's exact test.

## References

1. Wolf J, Johnston RB, Ambelu A, et al. Burden of disease attributable to unsafe drinking water, sanitation, and hygiene in domestic settings: a global analysis for selected adverse health outcomes. Lancet (London, England) 2023; 401(10393): 2060–71.

2. Troeger C, Blacker B, Khalil IA, et al. Estimates of the global, regional, and national morbidity, mortality, and aetiologies of lower respiratory infections in 195 countries, 1990–2016: a systematic analysis for the Global Burden of Disease Study 2016. The Lancet infectious diseases 2018; 18(11): 1191–210.

3. Estimates of global, regional, and national morbidity, mortality, and aetiologies of diarrhoeal diseases: a systematic analysis for the Global Burden of Disease Study 2015. The Lancet Infectious diseases 2017; 17(9): 909–48.

4. Berkman DS, Lescano AG, Gilman RH, Lopez SL, Black MM. Effects of stunting, diarrhoeal disease, and parasitic infection during infancy on cognition in late childhood: a follow-up study. Lancet (London, England) 2002; 359(9306): 564–71.

5. Walker SP, Chang SM, Powell CA, Simonoff E, Grantham-McGregor SM. Early childhood stunting is associated with poor psychological functioning in late adolescence and effects are reduced by psychosocial stimulation. J Nutr 2007; 137(11): 2464–9.

6. Tarleton JL, Haque R, Mondal D, Shu J, Farr BM, Petri WA, Jr. Cognitive effects of diarrhea, malnutrition, and Entamoeba histolytica infection on school age children in Dhaka, Bangladesh. The American journal of tropical medicine and hygiene 2006; 74(3): 475–81.

7. Leroy JL, Frongillo EA. Perspective: what does stunting really mean? A critical review of the evidence. Advances in Nutrition 2019; 10(2): 196–204.

8. WHO. Cholera in the WHO African Region: Weekly Regional Cholera Bulletin. 2024. https://www.afro.who.int/health-topics/disease-outbreaks/cholera-who-african-region#:~:text=As%20of%2031%20July%202024,(4%20375)%20deaths%20reported.

9. Moore SM, Azman AS, Zaitchik BF, et al. El Niño and the shifting geography of cholera in Africa. Proceedings of the National Academy of Sciences of the United States of America 2017; 114(17): 4436–41.

10. Quattrochi JP, Coville A, Mvukiyehe E, et al. Effects of a community-driven water, sanitation and hygiene intervention on water and sanitation infrastructure, access, behaviour, and governance: a cluster-randomised controlled trial in rural Democratic Republic of Congo. BMJ global health 2021; 6(5).

11. Bisimwa L, Williams C, Bisimwa JC, et al. Formative Research for the Development of Evidence-Based Targeted Water, Sanitation, and Hygiene Interventions to Reduce Cholera in Hotspots in the Democratic Republic of the Congo: Preventative Intervention for Cholera for 7 Days (PICHA7) Program. International journal of environmental research and public health 2022; 19(19).

12. George CM, Zohura F, Teman A, et al. Formative research for the design of a scalable water, sanitation, and hygiene mobile health program: CHoBI7 mobile health program. BMC public health 2019; 19(1): 1028.

13. George CM, Sanvura P, Bisimwa J-C, et al. Effects of a Water, Sanitation, and Hygiene Program on Diarrhea and Child Growth in the Democratic Republic of the Congo: A Cluster-Randomized Controlled Trial of the Preventative-Intervention-for-Cholera-for-7-Days (PICHA7) Program. 2024: 2024.12.16.24318942.

14. George CM, Monira S, Zohura F, et al. Effects of a Water, Sanitation and Hygiene Mobile Health Program on Diarrhea and Child Growth in Bangladesh: A Cluster-Randomized Controlled Trial of the CHoBI7 Mobile Health Program. Clinical infectious diseases : an official publication of the Infectious Diseases Society of America 2020.

15. George CM, Namunesha A, Endres K, et al. Epidemiologic and Genomic Surveillance of Vibrio cholerae and Effectiveness of Single-Dose Oral Cholera Vaccine, Democratic Republic of the Congo. Emerging infectious diseases 2025; 31(2): 288–97.

16. engageSPARK. engageSPARK platform. 2025. https://www.engagespark.com/.

17. Organization WH. Guidelines for drinking-water quality: incorporating the first and second addenda: World Health Organization; 2022.

18. Halder AK, Tronchet C, Akhter S, Bhuiya A, Johnston R, Luby SP. Observed hand cleanliness and other measures of handwashing behavior in rural Bangladesh. BMC public health 2010; 10: 545.

19. WHO/UNICEF. The JMP ladder for sanitation. 2020. https://washdata.org/monitoring/sanitation.

20. JMP/UNICEF. The JMP service ladder for drinking water. 2022.

21. Dupas P, Hoffmann V, Kremer M, Zwane AP. Targeting health subsidies through a nonprice mechanism: A randomized controlled trial in Kenya. Science (New York, NY) 2016; 353(6302): 889–95.

22. Dupas P, Nhlema B, Wagner Z, Wolf A, Wroe EJAEJEP. Expanding access to clean water for the rural poor: experimental evidence from Malawi. 2023; 15(1): 272–305.

23. Endres K, Mwishingo A, Thomas E, et al. A Quantitative and Qualitative Program Evaluation of a Case-Area Targeted Intervention to Reduce Cholera in Eastern Democratic Republic of the Congo. International journal of environmental research and public health 2023; 21(1).

